# The challenge of limited vaccine supplies: impact of prior infection on anti-spike IgG antibody trajectories after a single COVID-19 vaccination

**DOI:** 10.1101/2021.12.08.21267353

**Authors:** Jia Wei, Philippa C. Matthews, Nicole Stoesser, Ian Diamond, Ruth Studley, Emma Rourke, Duncan Cook, John I Bell, John N Newton, Jeremy Farrar, Alison Howarth, Brian D. Marsden, Sarah Hoosdally, E Yvonne Jones, David I Stuart, Derrick W. Crook, Tim E. A. Peto, A. Sarah Walker, David W. Eyre, Koen B. Pouwels, the COVID-19 Infection Survey team

## Abstract

Given high SARS-CoV-2 incidence, coupled with slow and inequitable vaccine roll-out, there is an urgent need for evidence to underpin optimum vaccine deployment, aiming to maximise global population immunity at speed. We evaluate whether a single vaccination in previously infected individuals generates similar initial and subsequent antibody responses to two vaccinations in those without prior infection. We compared anti-spike IgG antibody responses after a single dose of ChAdOx1, BNT162b2, or mRNA-1273 SARS-CoV-2 vaccines in the COVID-19 Infection Survey in the UK general population. In 100,849 adults who received at least one vaccination, 13,404 (13.3%) had serological and/or PCR evidence of prior infection. Prior infection significantly boosted antibody responses for all three vaccines, producing a higher peak level and longer half-life, and a response comparable to those without prior infection receiving two vaccinations. In those with prior infection, median time above the positivity threshold was estimated to last for >1 year after the first dose. Single-dose vaccination targeted to those previously infected may provide protection in populations with high rates of previous infection faced with limited vaccine supply, as an interim measure while vaccine campaigns are scaled up.

## Main

COVID-19 vaccines have moderate to high efficacy in preventing infections, severe illness, hospitalisation, and death, including the mRNA vaccines BNT162b2 and mRNA-1273, and adenovirus vaccine ChAdOx1^1–3^. By October 2021, more than 6 billion doses have been administered globally. However, 57% of the world’s population remain unvaccinated^4^, and coverage is highly uneven with low vaccination rates in many low-income countries^5^. For example, 58% of people in Europe and South America are fully vaccinated, 55% in North America, 49% in Asia and 7% in Africa^6^, with marked regional differences, for example 88% of the population in Portugal being fully vaccinated, versus 22% in Bosnia-Herzegovina. Previous infection also confers significant protection against re-infection^7,8^. However, prior infection rates vary widely, with high seroprevalences estimates in South Africa (48.5%), Ecuador (44.8%) and Peru (43.5%) versus 0.71% in Australia and New Zealand at various stages of the pandemic^9^.

Optimising global immunity and protection against infection is an urgent priority, to minimise deaths, morbidity, and socio-economic losses. While major initiatives seek to address inequalities in vaccine availability, including the COVAX Advance Market Commitment programme^10^, marked variation in global access persists. The WHO roadmap for prioritizing COVID-19 vaccine use in the context of limited access is appropriately focused on scale-up of equitable vaccine delivery^11^. Modelling studies suggest that prioritisation based on seropositivity substantially improves efficiency where seroprevalence is high^12^. Given this, a single-dose vaccine strategy for individuals with prior infection has already been adopted in some settings (e.g. Netherlands^13^, France, Italy, Germany^14^).

Understanding the extent to which prior infection influences antibody responses to vaccinations would inform a more consistent global approach to short-term interventions to optimise population immunity while vaccine deployment scales up. If a single vaccination invokes effective protection among those with prior infection, changing vaccine prioritisation as an interim measure may deliver higher population-level immunity faster, and may also make vaccination programs more affordable.

Existing studies have focused on how prior infection affects initial peak antibody responses^15^ or responses after two doses of vaccine^16^, showing prior infection significantly boosts vaccine-mediated antibody levels^17–20^. However, the durability of antibody response after a single vaccination is still unclear; whether a single dose can provide sustained effective protection for individuals with prior SARS-CoV-2 infection requires further investigation, particularly in terms of antibody waning after one dose of vaccination with prior infection versus two doses of vaccine without previous infection, which we have previously described^16^.

We used data from the United Kingdom’s national COVID-19 Infection Survey (ISRCTN21086382), to investigate the impact of prior infection on anti-trimeric spike IgG antibody responses following a single dose of ChAdOx1, BNT162b2, or mRNA-1273 vaccine, and compared the duration of protection for previously infected people receiving a single vaccination to previously uninfected people receiving two vaccinations. We included participants with at least one antibody measurement from 91 days before the first vaccination onwards up to the second vaccination (if received) or breakthrough infection post-first dose. From 8^th^ December 2020 to 18^th^ October 2021, 80,611 included participants received at least one dose of ChAdOx1 (10,168 [12.6%] with prior infection before the first vaccination), 56,024 at least one dose of BNT162b2 (9,556 [17.1%] with prior infection, reflecting many SARS-CoV-2 exposed healthcare workers receiving BNT162b2 early in the vaccination programme), and 3,545 at least one dose of mRNA-1273 (779 [22.0%] with prior infection, reflecting this vaccine being used later in the pandemic in younger age groups) (**Table S1**). The median age was 50 years (interquartile range IQR: 37-63). 75,000 (53.2%) were female, 130,542 (92.6%) reported white ethnicity, 1,214 (0.9%) black ethnicity, and 9,203 (6.5%) another ethnicity. 2,466 (1.7%) were healthcare workers, and 34,191 (24.3%) reported having a long-term health condition.

We modelled antibody trajectories using measurements from 28 days post-first dose for all participants (approximate peak levels, **Figure S1, S2**). We excluded participants who did not mount an anti-S antibody response to first vaccination (defined as all antibody measurements <16 BAU/mL, including ≥1 measurement ≥21 days after the first dose [similar to our previous studies^15,16^]): 4,940 (6.1%), 1,624 (2.9%), and 18 (0.5%) participants without prior infection, and 147 (1.8%), 48 (0.9%), 0 (0%) with prior infection with ChAdOx1, BNT162b2, and mRNA-1273, respectively.

59,469 participants with a single ChAdOx1 vaccination (7,211 (12.1%) previously infected) contributed 74,953 antibody measurements ≥28 days post-first dose, median (IQR) [range] 2 (1-2) [1-3] measurements per participant. Assuming antibody levels declined exponentially^16^, using multivariable Bayesian linear mixed models we estimated a median peak anti-spike IgG level of 449 BAU/mL (95% credible interval, Crl 434-464), and a median half-life of 85 days (95%Crl 76-96) for those with prior infection in the reference category (50 years, female, white ethnicity, not reporting a long-term health condition, not a healthcare worker, and deprivation percentile=60). This peak was substantially higher than our previous estimates after a second dose in participants without prior infection (160 BAU/mL [157-162] for the same reference category), and the half-life was similar (81 days [79-83])^16^. Those receiving one ChAdOx1 vaccination without prior infection had significantly lower peak levels, 84 BAU/mL (81-85), but no significant differences in half-life although slightly longer, 95 days (90-100) (**Figure 1a, Table S2**). **Table S3** compares peak levels and half-lives for different age groups, sex, and ethnicity. Previously infected participants with non-white ethnicity had higher post-first dose peak levels but slightly shorter half-lives (**Table 1, Figure S2**).

**Table 1.**
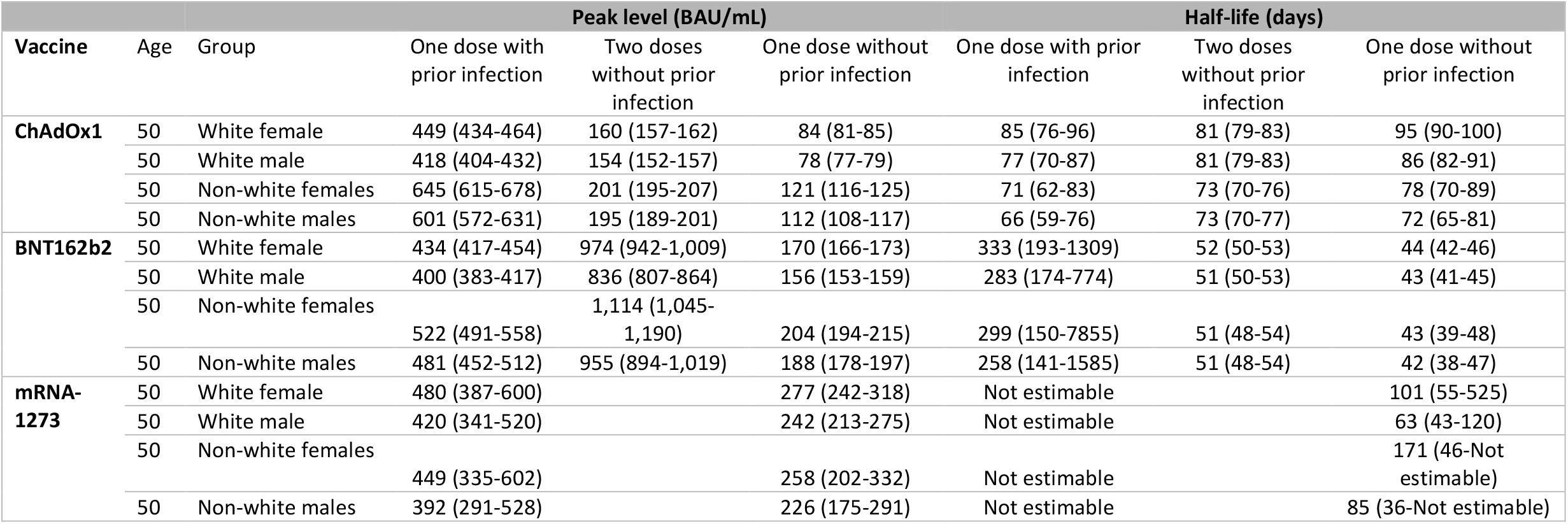
Posterior predicted median peak levels (BAU/mL) and half-lives (days) with 95% credible intervals in participants received one vaccination with prior infection, two vaccinations without prior infection, and one vaccination without prior infection, by vaccine type. Estimates for two vaccinations without prior infection were based on our previous analysis^16^. All estimates are at the reference age (50-year-old) and separated by sex (female vs male) and ethnicity (white vs non-white).

**Figure 1.**
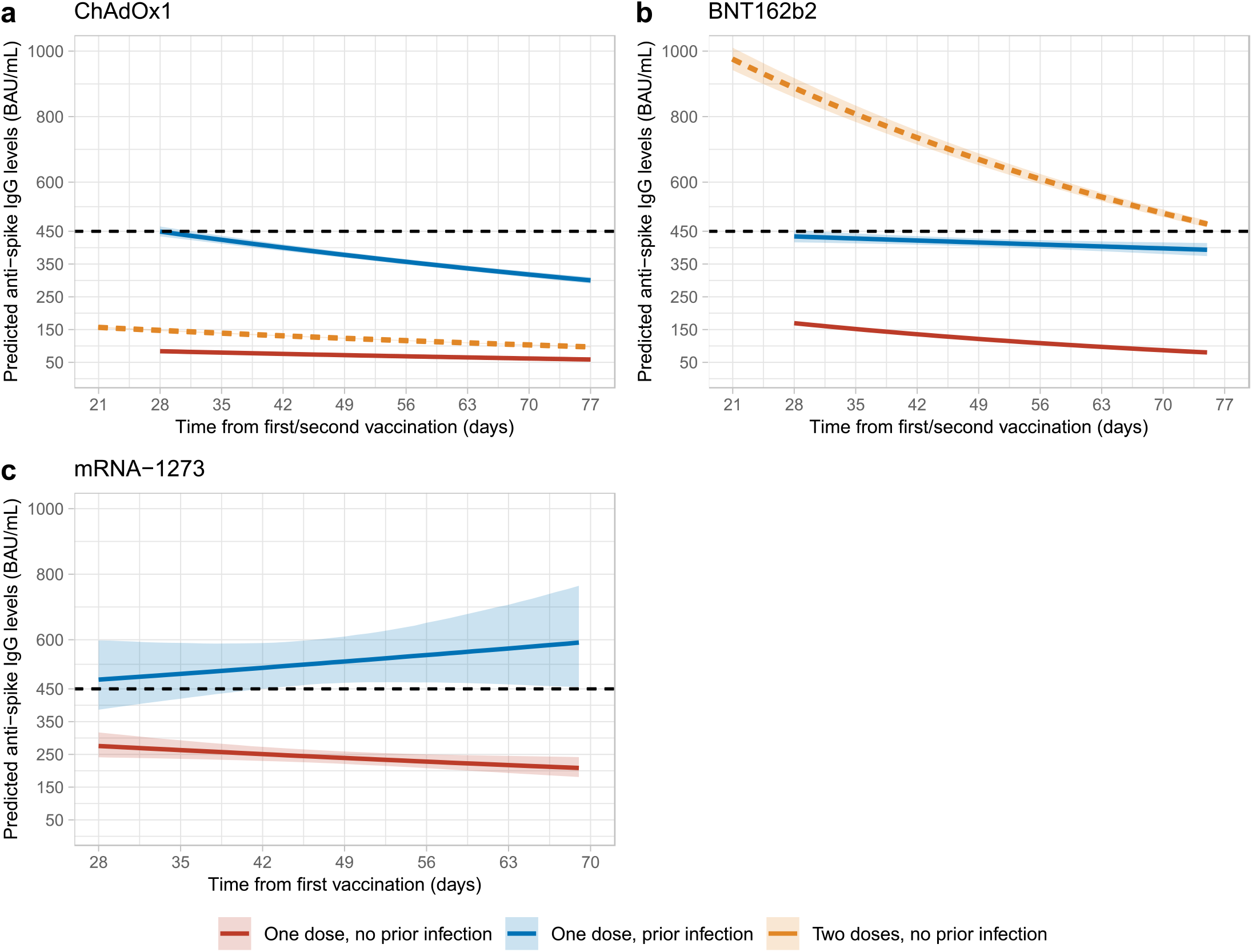
Posterior predicted trajectories of anti-spike IgG levels from 28 days post-first dose by prior infection status. **a**, 59,469 participants who received at least a single ChAdOx1 vaccination. **b**, 33,336 participants who received at least a single BNT162b2 vaccination. **c**, 2,443 participants who received at least a single mRNA-1273 vaccination. Plotted at reference categories: 50 years, female, white ethnicity, not reporting a long-term health condition, not a healthcare worker, and deprivation percentile=60. Black dotted line shows the upper quantification limit of 450 BAU/mL. Orange dotted lines in panel a and b were predicted trajectories starting from 21 days post-second dose for 92,584 and 51,034 participants who received two ChAdOx1 and BNT162b2 vaccination without prior infection reproduced from our previous analysis, plotted at the same reference categories^16^.

33,336 participants with a single BNT162b2 vaccination (4,803 (14.4%) previously infected) contributed 39,723 antibody measurements ≥28 days post-first dose, median (IQR) [range] 1 (1-2) [1-3] measurements per participant. For those with prior infection, the estimated median peak antibody level was 434 BAU/mL (95%Crl 417-454), and the half-life was 333 days (193-1309) at the reference category. The peak levels were lower than previously reported following two BNT162b2 vaccinations without prior infection (974 BAU/mL [942-1009]), but half-life was substantially longer (51 days [50-53])^16^. Similar to ChAdOx1, non-white ethnicity was associated with a higher peak level (**Table 1, Figure S2**). Those with one vaccination but no prior infection had lower peak levels (170 BAU/mL [166-173]) and shorter half-lives (44 days [42-46]) (**Figure 1b, Table S2**).

2,443 participants with one mRNA-1273 vaccination (364 (14.9%) previously infected) contributed 2,730 antibody measurements ≥21 days post-first dose, median (IQR) [range] 1 (1-2) [1-2] measurements per participant. For those receiving one vaccination with prior infection the estimated peak level was 480 BAU/mL (387-600) at the reference category, but the half-life was not estimable because antibody levels were not estimated to decline. No comparative data after two vaccinations were available, but participants without prior infection had lower peak levels after one vaccination (277 BAU/mL (242-318)) and half-lives was estimated to be 101 days (55-525) (**Table1, Figure 1c, Table S2**). mRNA-1273 elicited higher antibody levels (especially in those without prior infection) and had a lower percentage of seronegative non-responders versus BNT162b2 (0.5% vs 2.9%), consistent with findings after two doses^21^, and potentially explained by a higher spike protein delivery in mRNA-1273.

We estimated the duration of antibody positivity from first vaccination to levels falling to the antibody positivity threshold (23 BAU/mL, see Methods). For those with prior infection, the estimated median durations were 360-430 days for all age groups with ChAdOx1, and 650-1000 days for 20-40-year-olds with BNT162b2 following one vaccination. For mRNA-1273 and 60-80-year-olds with BNT162b2, antibody levels in some groups were also not estimated to decline and the upper credible intervals could not be defined for all groups (**Figure 2**). Females and those without long-term health conditions had longer estimated durations of seropositivity following all three vaccines. Conditional on seroconverting after one dose as described above, older participants had longer durations of seropositivity than younger participants following all three vaccines due to longer half-lives despite lower peak levels (**Table S2**). In our previous analysis of responses post-second vaccination, the time from the second dose to antibody levels falling to 23 BAU/mL was estimated to be around 250 days for those without prior infection who received two ChAdOx1 doses^16^, i.e., much shorter than those with prior infection receiving a single dose, regardless of age. For two BNT162b2 doses without prior infection, the estimated duration of seropositivity post second vaccination was 270-330 days. In this analysis, with previous infection and one vaccination, estimates were much longer, especially in the older age groups.

**Figure 2.**
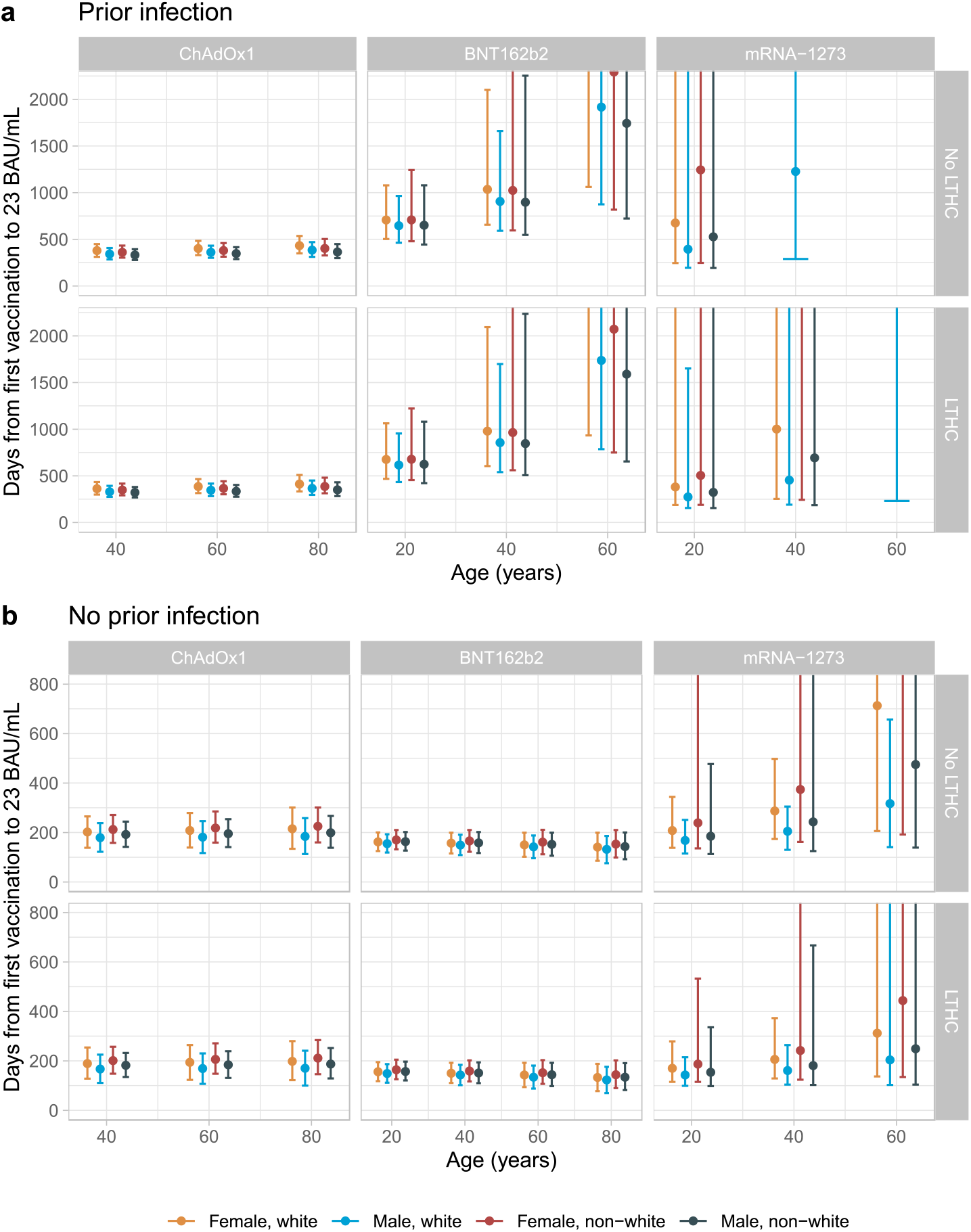
Posterior predicted days (95% credible interval) from the first vaccination to the positivity threshold of 23 BAU/mL in those with evidence of prior infection (panel a) and without evidence of prior infection (panel b). Estimates were separated by age, sex, ethnicity, long-term health condition (LTHC), and vaccine type. The y-axis is truncated at 2,000 days (panel a) for visualisation. For ChAdOx1, the 20-year-old group is not plotted because the vast majority of those receiving ChAdOx1 were ≥40 years. For mRNA-1273, the 80-year-old is not plotted because the vast majority of those receiving mRNA-1273 were ≤60 years. In panel a, 80-year-old in BNT162b2, some 40- and 60-year-old groups in mRNA-1273 are not plotted because their antibody levels were not estimated to decline so no duration could be estimated. Equivalent estimates after second vaccination are provided in^16^.

We previously reported antibody levels correlating with protection from infection, providing context to the observed antibody levels; post-vaccine levels of 107 BAU/mL for ChAdOx1 and 94 BAU/mL for BNT162b2 are associated with 67% protection against new infection in those without prior infection, compared with 33 BAU/mL in those unvaccinated with prior infection^16^. Data were insufficient to estimate correlates of protection for those with prior infection^16^, but, since levels associated with the same degree of protection were lower for unvaccinated individuals, if we conservatively assume the threshold levels are similar post any vaccination, the duration providing >67% protection is estimated to be around 170-220 days for a single ChAdOX1 vaccination, and over a year for a single BNT162b2 vaccination in those with prior infection. Since the duration providing >67% protection in unvaccinated individuals with natural infection was estimated to be 1-2 years^16,22^, it is highly likely that the duration of protection is >1 year for those with prior infection receiving a single vaccination. In those without prior infection, using these thresholds, a single ChAdOx1 vaccination would not reach the required antibody level, while a single BNT162b2 vaccination would provide 50-100 days of protection for people <60 years (**Figure S3**).

Higher antibody levels post SARS-CoV-2 vaccination in previously infected individuals have been reported^17–20^, but these studies did not estimate the trajectory of antibody response. We found that in those with prior infection, not only were antibody peak levels higher in all three vaccines, by 200-400 BAU/mL, but the subsequent waning was also slower in BNT162b2, supporting sustained protection from a single dose in previously infected individuals. The combination of prior infection with a single vaccination resulted in similar antibody levels regardless of vaccine type (**Figure 1**), despite single ChAdOx1 vaccination resulting in lower peak levels than BNT162b2 in those without prior infection.

Our results could help inform and optimise global immunisation strategies, deploying limited resources in the most effective way to deliver maximum population immunity at speed during a period when incidence remains high and vaccine access is not yet universal. Given the low percentage of vaccinated individuals and high SARS-CoV-2 seroprevalence in some settings^23–25^, assuming widespread prior infection, a single vaccination could optimise population-level protection as a short-term measure.

However, such a blanket approach does result in infection-naive individuals initially receiving one vaccination only, which provides suboptimal protection^8^. To reduce this risk, where previous infection is expected to be high, another option would be to stratify individual vaccination based on an affordable and rapid lateral flow immunoassay (LFIA) for antibody detection, and information on previous PCR positivity. An extensive comparison of different fingerprick-based LFIA antibody tests reported high specificity (98.8–99.8%)^26^, meaning that there would be few false positives in a high prevalence setting. Sensitivity was lower (69%-86%)^26^, which would reduce efficiency, but not vaccine programme effectiveness. Although exact prices will vary by manufacturer and may be negotiated, the cost can be as low as US$1-2 per test^27^. In comparison, a ChAdOx1 vaccine costs around $4 per dose excluding delivery and storage costs^28^.

For an unvaccinated population of 10 million individuals willing to be vaccinated with a true seroprevalence of 50%, a LFIA with 99% specificity and 80% sensitivity costing $1.5 per test, and a cost of $5 per vaccination (including delivery costs and storage), an LFIA test performed at the first vaccination visit would correctly identify 4,050,000 individuals as having antibodies and eligible for a one-dose schedule – of which 50,000 would be incorrectly identified as having antibodies – the remaining 5,950,000 being invited for a second vaccination according to the agreed dosing interval. This would result in 4.05 million fewer vaccinations needed in the short-term to produce equivalent population immunity to a universal two-dose campaign. This approach delivers a cost saving of $5.25 million which could be re-invested in securing robust long-term vaccine access (for results with other settings regarding LFIA test sensitivity and specificity, population size, true antibody prevalence, costs per vaccination and LFIA, see https://herc.shinyapps.io/Serology_vaccine_prioritisation/).

Study limitations include that we did not measure other immune responses, including T cell or innate immune responses. We were also unable to fully estimate rates of waning following mRNA-1273, reflecting the fact that 41% of antibody levels were above the upper limit of quantification for our assay (450 BAU/mL) and sample size was relatively small. We only measured anti-spike IgG antibody levels using a single assay; however, we calibrated antibody levels to WHO BAU/mL units for comparison with other studies; assay results have been previously shown to correlate closely with neutralising activity^16^. Most of our participants reported white ethnicity (92.6%), so wider generalizability to non-white ethnic groups is less well defined and our data were insufficient to model other ethnic groups separately. However, estimated durations of protection for non-white participants were broadly as long or longer than for white participants, with non-white ethnicity associated with higher peak levels after a single ChAdOX1 or BNT162b2 vaccination.

Data are still awaited to determine the impact of the recent emergence of the Omicron variant, and the relationship between prior infection/vaccination and immunological protection from this and other new variants. For this reason, all approaches to vaccine scheduling will need to remain under intense scrutiny, and the international focus must remain firmly on assuring equitable access to full vaccination in all population settings, which will also mitigate the potential for further variants of concern to emerge.

In summary, prior infection significantly boosts antibody responses after a single ChAdOx1, BNT162b2, or mRNA-1273 vaccination, producing higher peak levels and longer half-lives, comparable or even better to that obtained from two vaccinations in those without prior infection. Based on the positivity threshold and previously reported correlates of protection, those with prior infection could be protected from infection for >1 year after a single vaccination. While recent studies show that a third vaccination boosts antibody responses^29^ and provides better protection against infection than two vaccinations^30^, and two vaccinations plus prior infection provides better protection against re-infection than prior infection alone^8^, a large part of the global population did not yet have the chance to get their first vaccination due to limited vaccine supplies. These results could inform vaccine strategies, providing an evidence-base to optimise population immunity in the context of resource limitations, while international SARS-CoV-2 immunisation programmes are scaled up and secured.

## Online Methods

### Population and survey

The UK’s Office for National Statistics (ONS) COVID-19 Infection Survey (CIS) (ISRCTN21086382) randomly and continuously recruited private households to provide a representative sample across its four countries (England, Wales, Northern Ireland, Scotland). At the first visit, participants were asked for consent for optional follow-up visits every week for the next month, then monthly for 12 months or to April 2022. Written informed consent was taken from individuals ≥2 years (children aged <2 years were not eligible for the study). For those 2-15 years this consent was obtained from parents/carers, while those 10-15 years also provided written assent.

Socio-demographic characteristics, behaviours, and vaccination data were collected. Combined nose and throat swabs were taken from all consenting participants for SARS-CoV-2 PCR testing. Blood samples were taken from individuals ≥16 years from 10-20% randomly selected households monthly for serological testing, and participants who tested swab positive and their household members were also invited to provide blood samples at follow-up visits. Details on the sampling design are provided elsewhere^31^. From April 2021, additional participants were invited to provide blood samples monthly to assess vaccine responses. The study protocol is available at https://www.ndm.ox.ac.uk/covid-19/covid-19-infection-survey/protocol-and-information-sheets. The study received ethical approval from the South Central Berkshire B Research Ethics Committee (20/SC/0195).

### Vaccination data

Self-reported vaccinations were obtained from participants at visits, including vaccination type, number of doses, and vaccination dates. Participants from England were also linked to the National Immunisation Management Service (NIMS), which contains all individuals’ vaccination data in the English National Health Service COVID-19 vaccination programme. There was good agreement between self-reported and administrative vaccination data (98% on type and 95% on date^32^). We used vaccination data from NIMS where available for participants from England, and otherwise data from the survey.

### Laboratory testing

Combined nose and throat swabs were tested at high-throughput national “Lighthouse” laboratories in Glasgow (from 16 August 2020 to present) and Milton Keynes (from 26 April 2020 to 8 February 2021). PCR outputs were analysed using UgenTec Fast Finder 3.300.5 (TaqMan 2019-nCoV Assay Kit V2 UK NHS ABI 7500 v2.1; UgenTec), with an assay-specific algorithm and decision mechanism that allows conversion of amplification assay raw data into test results with minimal manual intervention. Positive samples are defined as having at least a single N and/or ORF1ab gene detected, and PCR traces exhibited an appropriate morphology. The S gene alone is not considered to be positive^31^. SARS-CoV-2 antibody levels were tested on venous or capillary blood samples using an ELISA detecting anti-trimeric spike IgG developed by the University of Oxford^31,33^. Normalised results are reported in ng/ml of mAb45 monoclonal antibody equivalents. Before 26 February 2021, the assay used fluorescence detection as previously described, with a positivity threshold of 8 million units validated on banks of known SARS-CoV-2 positive and negative samples^33^. After this, it used a commercialised CE-marked version of the assay, the Thermo Fisher OmniPATH 384 Combi SARS-CoV-2 IgG ELISA (Thermo Fisher Scientific), with the same antigen and colorimetric detection. mAb45 is the manufacturer-provided monoclonal antibody calibrant for this quantitative assay. To allow conversion of fluorometrically determined values in arbitrary units, we compared 3,840 samples which were run in parallel on both systems. A piece-wise linear regression was used to generate the following conversion formula:

(1) log_10_(mAb45 units) = 0.221738 + 1.751889e-07*fluorescence_units + 5.416675e-07*(fluorescence_units>9190310)*(fluorescence_units-9190310)

We calibrated the results of the Thermo Fisher OmniPATH assay into WHO international units (binding antibody unit, BAU/mL) using serial dilutions of National Institute for Biological Standards and Control (NIBSC) Working Standard 21/234. The NIBSC 21/234 Working Standard has been previously calibrated against the WHO International Standard for anti-SARS-CoV-2 immunoglobulin (NIBSC code 20/136), with anti-spike IgG potency of 832 BAU/mL (95%CI 746-929). We generated 2-fold dilutions of 21/234 between 1:400 and 1:8000 from three separate batches on three separate days. Results from a total of 63 diluted samples were merged and a linear regression model fitted constrained to have an intercept of zero to convert mAB45 units in ng/ml for samples diluted at 1:50 to BAU/mL:

BAU/mL = 0.559 * [mAb45 concentration in ng/mL at 1:50]

23 BAU/mL was used as the threshold for an IgG positive or negative result (corresponding to the 8 million units with fluorescence detection). Given the lower and upper limits of the assay, measurements <1 BAU/mL (2533 observations, 0.8%) and >450 BAU/mL (28,086 observations, 8.4%) were truncated at 1 and 450 BAU/mL, respectively.

### Statistical analysis

For the current study, participants aged ≥16 years who received at least a single vaccination with ChAdOx1 or BNT162b2 or mRNA-1273 with antibody measurements from 8^th^ December 2020 until 18^th^ October 2021 were included. Participants with prior infection (before vaccination) were defined as 1) having a positive PCR swab test in the survey or the linked English national testing programme; 2) having a positive anti-spike IgG result (≥23 BAU/mL) before vaccination; 3) having two consecutive positive anti-nucleocapsid IgG results (≥17 BAU/mL); or 4) self-reporting a positive swab test in the survey. The infection date was defined as the earliest recorded date from the above definitions. Age was truncated at 85 years to reduce the influence of outliers.

To estimate antibody waning, we excluded a small number of participants who were considered as non-responders after the first dose, defined as all antibody measurements being <16 BAU/mL and having at least one antibody measurement 21 days after the first dose (N=5,087 excluded for ChAdOx1, N=1,672 excluded for BNT162b2, N=18 excluded for mRNA-1273).

Bayesian linear mixed interval-censored models were used to estimate changes in antibody levels after the first vaccination with ChAdOX1, BNT162b2, or mRNA-1273. Antibody measurements taken after the second vaccination or after infection that happened post-first dose were excluded. We included antibody measurements from 28 days post-first dose to reflect the peak level for participants <60 years. We excluded measurements taken after the 90^th^ percentile of the observed time points to avoid outlier influence (77, 75, and 69 days post-first dose for ChAdOx1, BNT162b2, and mRNA-1273).

We used a multivariable model to examine the association between peak levels and antibody half-lives with continuous age (16-85 years), sex, ethnicity (white vs non-white), reporting having a long-term health condition, reporting working in patient-facing healthcare, deprivation percentile, and prior infection status. We assumed an exponential fall in antibody levels over time, i.e., a linear decline on a log2 scale^16^. Population-level fixed effects, individual-level random effects for intercept and slope, and correlation between random effects were included in the models. The outcome was right-censored at 450 BAU/mL reflecting truncation of IgG values at the upper limit of quantification (i.e. all measurements truncated to 450 BAU/mL were considered to be >450 BAU/mL in analyses). For each model, weakly informative priors were used. Four chains were run per model with 4,000 iterations and a warm-up period of 2,000 iterations to ensure convergence, which was confirmed visually and by ensuring the Gelman-Rubin statistic was <1.05. 95% credible intervals were calculated using highest posterior density intervals.

All analyses were performed in R 4.1 using the following packages: tidyverse (version 1.3.1), brms (version 2.15.0), arsenal (version 3.4.0), cowplot (version 1.1.1), bayesplot (version 1.8.1), and tidybayes (version 3.0.1).

## Data Availability

Data are still being collected for the COVID-19 Infection Survey. De-identified study data are available for access by accredited researchers in the ONS Secure Research Service (SRS) for accredited research purposes under part 5, chapter 5 of the Digital Economy Act 2017. For further information about accreditation, contact or visit the SRS website.

https://www.ons.gov.uk/aboutus/whatwedo/statistics/requestingstatistics/approvedresearcherscheme

## Acknowledgements

We are grateful for the support of all COVID-19 Infection Survey participants.

This study is funded by the Department of Health and Social Care with in-kind support from the Welsh Government, the Department of Health on behalf of the Northern Ireland Government and the Scottish Government. JW is supported by University of Oxford and the China Scholarship Council. ASW, TEAP, NS, DE, KBP are supported by the National Institute for Health Research (NIHR) Health Protection Research Unit in Healthcare Associated Infections and Antimicrobial Resistance (NIHR200915), a partnership between the UK Health Security Agency (UKHSA) and the University of Oxford. ASW and TEAP are also supported by the NIHR Oxford Biomedical Research Centre. KBP is also supported by the Huo Family Foundation. ASW is also supported by core support from the Medical Research Council UK to the MRC Clinical Trials Unit [MC_UU_12023/22] and is an NIHR Senior Investigator. PCM is funded by Wellcome (intermediate fellowship, grant ref 110110/Z/15/Z) and holds an NIHR Oxford BRC Senior Fellowship award. DWE is supported by a Robertson Fellowship and an NIHR Oxford BRC Senior Fellowship. NS is an Oxford Martin Fellow and holds an NIHR Oxford BRC Senior Fellowship. The views expressed are those of the authors and not necessarily those of the National Health Service, NIHR, Department of Health and Social Care, or UKHSA.

For the purpose of open access, the authors have applied a Creative Commons Attribution (CC BY) licence to any Author Accepted Manuscript version arising.

## COVID-19 Infection Survey team group authorship

**Office for National Statistics**: Sir Ian Diamond, Emma Rourke, Ruth Studley, Tina Thomas, Duncan Cook.

**Office for National Statistics COVID Infection Survey Analysis and Operations teams**, in particular Daniel Ayoubkhani, Russell Black, Antonio Felton, Megan Crees, Joel Jones, Lina Lloyd, Esther Sutherland.

**University of Oxford, Nuffield Department of Medicine**: Ann Sarah Walker, Derrick Crook, Philippa C Matthews, Tim Peto, Emma Pritchard, Nicole Stoesser, Karina-Doris Vihta, Jia Wei, Alison Howarth, George Doherty, James Kavanagh, Kevin K Chau, Stephanie B Hatch, Daniel Ebner, Lucas Martins Ferreira, Thomas Christott, Brian D Marsden, Wanwisa Dejnirattisai, Juthathip Mongkolsapaya, Sarah Cameron, Phoebe Tamblin-Hopper, Magda Wolna, Rachael Brown, Sarah Hoosdally, Richard Cornall, David I Stuart, Gavin Screaton.

**University of Oxford, Nuffield Department of Population Health**: Koen Pouwels.

**University of Oxford, Big Data Institute:** David W Eyre, Katrina Lythgoe, David Bonsall, Tanya Golubchik, Helen Fryer.

**University of Oxford, Radcliffe Department of Medicine**: John Bell.

**Oxford University Hospitals NHS Foundation Trust:** Stuart Cox, Kevin Paddon, Tim James.

**University of Manchester**: Thomas House.

**Public Health England**: John Newton, Julie Robotham, Paul Birrell.

**IQVIA**: Helena Jordan, Tim Sheppard, Graham Athey, Dan Moody, Leigh Curry, Pamela Brereton.

**National Biocentre**: Ian Jarvis, Anna Godsmark, George Morris, Bobby Mallick, Phil Eeles.

**Glasgow Lighthouse Laboratory**: Jodie Hay, Harper VanSteenhouse.

**Department of Health and Social Care**: Jessica Lee.

**Welsh Government**: Sean White, Tim Evans, Lisa Bloemberg.

**Scottish Government**: Katie Allison, Anouska Pandya, Sophie Davis.

**Public Health Scotland**: David I Conway, Margaret MacLeod, Chris Cunningham.

## Author Contributions

The study was designed and planned by ASW, JF, JB, JN, ID and KBP, and is being conducted by ASW, RS, DC, ER, AH, BM, TEAP, PCM, NS, SH, EYJ, DIS, DWC and DWE. This specific analysis was designed by JW, DWE, ASW, and KBP. JW contributed to the statistical analysis of the survey data. JW, DWE, KBP and ASW drafted the manuscript and all authors contributed to interpretation of the data and results and revised the manuscript. All authors approved the final version of the manuscript.

## Code availability

A copy of the analysis code is available at https://github.com/jiaweioxford/COVID19_antibody_response_first_dose.

