## Supplementary Figures and Tables for "The challenge of limited vaccine supplies: impact of prior infection on anti-spike IgG antibody trajectories after a single COVID-19 vaccination"

| | Without prior infection | | | With prior infection | | | | |
|---|---|---|---|---|---|---|---|---|
| | ChAdOx1 (N=70,443) | BNT162b2 (N=46,468) | mRNA-1273 (N=3,545) | ChAdOx1 (N=10,168) | BNT162b2 (N=9,556) | mRNA-1273 (N=779) | Total (N=140,959) | p value |
| **Age** | | | | | | | | < 0.001 |
| **Median** | 55 | 39 | 35 | 52 | 34 | 32 | 50 | |
| **Q1, Q3** | 46, 64 | 30, 64 | 28, 41 | 44, 61 | 25, 49 | 26, 39 | 37, 63 | |
| **Sex** | | | | | | | | < 0.001 |
| **Female** | 37,239 (52.9%) | 25,128 (54.1%) | 1,719 (48.5%) | 5,404 (53.1%) | 5,150 (53.9%) | 360 (46.2%) | 75,000 (53.2%) | |
| **Male** | 33,204 (47.1%) | 21,340 (45.9%) | 1,826 (51.5%) | 4,764 (46.9%) | 4,406 (46.1%) | 419 (53.8%) | 65,959 (46.8%) | |
| **Ethnicity** | | | | | | | | < 0.001 |
| **White** | 66,057 (93.8%) | 42,871 (92.3%) | 3,270 (92.2%) | 9,297 (91.4%) | 8,383 (87.7%) | 664 (85.2%) | 130,542 (92.6%) | |
| **Black** | 560 (0.8%) | 359 (0.8%) | 18 (0.5%) | 122 (1.2%) | 145 (1.5%) | 10 (1.3%) | 1,214 (0.9%) | |
| **Others** | 3,826 (5.4%) | 3,238 (7.0%) | 257 (7.2%) | 749 (7.4%) | 1,028 (10.8%) | 105 (13.5%) | 9,203 (6.5%) | |
| **Household size** | | | | | | | | < 0.001 |
| **1** | 12,760 (18.1%) | 7,310 (15.7%) | 453 (12.8%) | 1,641 (16.1%) | 1,162 (12.2%) | 88 (11.3%) | 23,414 (16.6%) | |
| **2** | 32,576 (46.2%) | 21,092 (45.4%) | 1,381 (39.0%) | 4,171 (41.0%) | 3,606 (37.7%) | 298 (38.3%) | 63,124 (44.8%) | |
| **3** | 11,429 (16.2%) | 8,183 (17.6%) | 750 (21.2%) | 1,826 (18.0%) | 1,999 (20.9%) | 155 (19.9%) | 24,342 (17.3%) | |
| **4** | 10,100 (14.3%) | 6,871 (14.8%) | 718 (20.3%) | 1,807 (17.8%) | 1,851 (19.4%) | 164 (21.1%) | 21,511 (15.3%) | |
| **5+** | 3,578 (5.1%) | 3,012 (6.5%) | 243 (6.9%) | 723 (7.1%) | 938 (9.8%) | 74 (9.5%) | 8,568 (6.1%) | |
| **Deprivation percentile** | | | | | | | | < 0.001 |
| **Median** | 62 | 60 | 58 | 59 | 56 | 55 | 61 | |
| **Q1, Q3** | 38, 82 | 37, 81 | 35, 81 | 34, 81 | 31, 78 | 29, 79 | 37, 81 | |
| **Region** | | | | | | | | < 0.001 |
| **North East** | 2,999 (4.3%) | 1,771 (3.8%) | 106 (3.0%) | 447 (4.4%) | 403 (4.2%) | 30 (3.9%) | 5,756 (4.1%) | |
| **North West** | 7,257 (10.3%) | 4,839 (10.4%) | 316 (8.9%) | 1,488 (14.6%) | 1,306 (13.7%) | 101 (13.0%) | 15,307 (10.9%) | |
| **Yorkshire and the Humber** | 6,035 (8.6%) | 3,579 (7.7%) | 418 (11.8%) | 929 (9.1%) | 797 (8.3%) | 96 (12.3%) | 11,854 (8.4%) | |
| **East Midlands** | 4,809 (6.8%) | 3,142 (6.8%) | 120 (3.4%) | 701 (6.9%) | 609 (6.4%) | 19 (2.4%) | 9,400 (6.7%) | |
| **West Midlands** | 5,812 (8.3%) | 3,637 (7.8%) | 189 (5.3%) | 852 (8.4%) | 737 (7.7%) | 32 (4.1%) | 11,259 (8.0%) | |
| **East of England** | 6,644 (9.4%) | 4,287 (9.2%) | 341 (9.6%) | 954 (9.4%) | 813 (8.5%) | 57 (7.3%) | 13,096 (9.3%) | |
| **London** | 8,519 (12.1%) | 7,804 (16.8%) | 755 (21.3%) | 2,000 (19.7%) | 2,361 (24.7%) | 227 (29.1%) | 21,666 (15.4%) | |

| | | | | | | | | |
|---|---|---|---|---|---|---|---|---|
| **South East** | 10,220 (14.5%) | 5,827 (12.5%) | 564 (15.9%) | 1,215 (11.9%) | 922 (9.6%) | 109 (14.0%) | 18,857 (13.4%) | |
| **South West** | 6,142 (8.7%) | 4,017 (8.6%) | 311 (8.8%) | 509 (5.0%) | 498 (5.2%) | 42 (5.4%) | 11,519 (8.2%) | |
| **Northern Ireland** | 1,912 (2.7%) | 1,781 (3.8%) | 15 (0.4%) | 189 (1.9%) | 268 (2.8%) | 4 (0.5%) | 4,169 (3.0%) | |
| **Scotland** | 6,061 (8.6%) | 3,818 (8.2%) | 333 (9.4%) | 521 (5.1%) | 509 (5.3%) | 56 (7.2%) | 11,298 (8.0%) | |
| **Wales** | 4,033 (5.7%) | 1,966 (4.2%) | 77 (2.2%) | 363 (3.6%) | 333 (3.5%) | 6 (0.8%) | 6,778 (4.8%) | |
| **Report working in patient-facing healthcare** | | | | | | | | < 0.001 |
| **No** | 69,773 (99.0%) | 45,224 (97.3%) | 3,518 (99.2%) | 9,995 (98.3%) | 9,207 (96.3%) | 776 (99.6%) | 138,493 (98.3%) | |
| **Yes** | 670 (1.0%) | 1,244 (2.7%) | 27 (0.8%) | 173 (1.7%) | 349 (3.7%) | 3 (0.4%) | 2,466 (1.7%) | |
| **Report having a long-term health condition** | | | | | | | | < 0.001 |
| **No** | 52,145 (74.0%) | 35,245 (75.8%) | 3,093 (87.2%) | 7,766 (76.4%) | 7,826 (81.9%) | 693 (89.0%) | 106,768 (75.7%) | |
| **Yes** | 18,298 (26.0%) | 11,223 (24.2%) | 452 (12.8%) | 2,402 (23.6%) | 1,730 (18.1%) | 86 (11.0%) | 34,191 (24.3%) | |

**Table S1. Characteristics of participants with a single ChAdOx1, BNT162b2, or mRNA-1273 vaccination and at least one antibody measurement from 91 days before the first vaccination up to the second vaccination (if received) or breakthrough infections post-first vaccination.**

| | | ChAdOx1 | | | BNT162b2 | | | mRNA-1273 | | |
|---|---|---|---|---|---|---|---|---|---|---|
| **Adjusted baseline** | Peak level (Intercept) | 84 | 81 | 85 | 170 | 164 | 173 | 277 | 242 | 318 |
| | IgG half-life (slope) | 95 | 90 | 100 | 44 | 42 | 46 | 101 | 55 | 525 |
| **Prior infection** | Peak level: No | | | | | | | | | |
| | IgG half-life: No | | | | | | | | | |
| | Change in peak level: Yes | **365** | **351** | **380** | **265** | **248** | **283** | **203** | **122** | **308** |
| | Change in half-life: Yes | -10 | -19 | 1 | **289** | **147** | **1155** | | **Not estimable** | |
| **Age** | Peak level: 50 years | | | | | | | | | |
| | IgG half-life: 50 years | | | | | | | | | |
| | Change in peak level: per 10-years older | **-4** | **-5** | **-4** | **-28** | **-29** | **-27** | **-58** | **-69** | **-45** |
| | Change in half-life: per 10-years older | **6** | **3** | **9** | **3** | **2** | **4** | | Not estimable | |
| **Sex** | Peak level: Female | | | | | | | | | |
| | IgG half-life: Female | | | | | | | | | |
| | Change in peak level: Male | **-6** | **-7** | **-4** | **-14** | **-18** | **-9** | **-35** | **-68** | **-4** |
| | Change in half-life: Male | **-9** | **-15** | **-3** | -1 | -3 | 1 | -35 | -346 | 14 |
| **Ethnicity** | Peak level: White | | | | | | | | | |
| | IgG half-life: White | | | | | | | | | |
| | Change in peak level: Non-white | **37** | **32** | **41** | **35** | **24** | **45** | -18 | -74 | 47 |
| | Change in half-life: Non-white | **-16** | **-26** | **-5** | -1 | -5 | 5 | 70 | -9 | Not estimable |
| **Report having a long-term health condition** | Peak level: No | | | | | | | | | |
| | IgG half-life: No | | | | | | | | | |
| | Change in peak level: Yes | **-6** | **-7** | **-4** | **-17** | **-22** | **-13** | 1 | -44 | 52 |
| | Change in half-life: Yes | -3 | -10 | 5 | 0 | -3 | 3 | -42 | -348 | 58 |
| **Report working in patient-facing healthcare** | Peak level: No | | | | | | | | | |
| | IgG half-life: No | | | | | | | | | |
| | Change in peak level: Yes | 8 | 0 | 18 | -12 | -25 | 1 | -71 | -187 | 164 |
| | Change in half-life: Yes | -21 | -38 | 6 | **24** | **11** | **48** | | Not estimable | |
| **Deprivation percentile** | Peak level: 60 (median) | | | | | | | | | |
| | IgG half-life: 60 (median) | | | | | | | | | |

| | | | | | | | | | | |
|---|---|---|---|---|---|---|---|---|---|---|
| | Change in peak level: per 10 percentile higher | 0 | -1 | 0 | 0 | -1 | 1 | 4 | -2 | 10 |
| | Change in half-life: per 10 percentile higher | 1 | -1 | 2 | 0 | 0 | 1 | -3 | -102 | 23 |

**Table S2. Posterior median and 95% credible intervals for anti-spike IgG peak level (intercept) (BAU/mL) and half-life (slope) (days) in the multivariable models in 59,469 participants with first ChAdOx1 vaccination, 33,336 participants with first BNT162b2 vaccination, and 2,443 participants with first mRNA-1273 vaccination**. The reference categories in the multivariable model are: no prior infection, 50-year-old, female, white ethnicity, not reporting a long-term health condition, not a healthcare worker, and deprivation percentile=60. Bold numbers indicate an effect not compatible with chance (95% credible interval excludes no effect). Model results are used to calculate estimations in **Table 1** and **Table S3**. In mRNA-1273, some numbers are not estimable because antibody responses in certain groups were not estimated to decline.

| | | | Peak level (BAU/mL) | | | Half-life (days) | | | Time to positivity threshold 23 BAU/mL (days) | | |
|---|---|---|---|---|---|---|---|---|---|---|---|
| | | | One dose with prior infection | Two doses without prior infection | One dose without prior infection | One dose with prior infection | Two doses without prior infection | One dose without prior infection | One dose with prior infection | Two doses without prior infection | One dose without prior infection |
| **ChAd Ox1** | 40 | White female | 474 (458-490) | 157 (154-159) | 89 (87-90) | 80 (72-91) | 81 (79-83) | 89 (84-96) | 379 (312-449) | 246 (170-322) | 202 (138-265) |
| | | White male | 441 (426-457) | 152 (150-154) | 82 (81-84) | 74 (67-83) | 82 (80-84) | 82 (77-87) | 342 (284-407) | 244 (167-318) | 179 (122-238) |
| | | Non-white female | 681 (648-715) | 197 (192-203) | 127 (122-132) | 68 (60-79) | 73 (70-77) | 75 (67-85) | 361 (303-433) | 248 (181-318) | 212 (158-271) |
| | | Non-white male | 633 (603-666) | 191 (186-197) | 118 (114-123) | 64 (57-73) | 74 (70-77) | 69 (62-78) | 332 (278-394) | 246 (176-316) | 192 (142-244) |
| | 60 | White female | 426 (411-440) | 162 (160-165) | 80 (78-81) | 89 (79-101) | 80 (78-82) | 100 (95-107) | 401 (330-483) | 246 (169-323) | 208 (139-279) |
| | | White male | 396 (383-410) | 157 (155-160) | 74 (73-75) | 81 (73-92) | 80 (78-82) | 91 (86-96) | 361 (301-432) | 244 (169-318) | 181 (117-246) |
| | | Non-white female | 612 (582-643) | 204 (198-211) | 114 (110-119) | 74 (65-87) | 72 (69-76) | 82 (73-95) | 380 (314-460) | 248 (182-317) | 218 (159-285) |
| | | Non-white male | 569 (542-598) | 198 (192-204) | 106 (102-111) | 69 (61-80) | 73 (69-76) | 75 (67-86) | 347 (288-416) | 245 (178-314) | 195 (141-254) |
| | 80 | White female | 383 (368-398) | 168 (164-172) | 72 (70-73) | 100 (86-118) | 79 (76-81) | 114 (103-128) | 432 (349-536) | 247 (174-321) | 215 (134-301) |
| | | White male | 356 (342-371) | 163 (159-166) | 67 (65-68) | 90 (79-105) | 79 (76-82) | 102 (93-113) | 384 (312-470) | 243 (173-318) | 184 (113-258) |
| | | Non-white female | 550 (520-581) | 212 (204-219) | 103 (98-108) | 82 (69-100) | 71 (68-75) | 91 (79-110) | 402 (327-503) | 248 (181-315) | 225 (160-301) |
| | | Non-white male | 512 (484-541) | 205 (198-212) | 96 (91-100) | 75 (65-90) | 71 (68-75) | 83 (72-98) | 364 (298-449 | 247 (178-316) | 199 (138-267) |
| **BNT16 2b2** | 20 | White female | 751 (718-787) | 1243 (1196-1293) | 293 (285-302) | 135 (101-203) | 51 (50-53) | 37 (35-39) | 707 (503-1078) | 323 (238-405) | 162 (125-200) |
| | | White male | 691 (661-725) | 1066 (1024-1110) | 270 (263-278) | 126 (96-183) | 51 (49-53) | 36 (34-38) | 646 (463-965) | 308 (223-392) | 155 (119-193) |
| | | Non-white female | 904 (850-964) | 1421 (1331-1520) | 353 (335-372) | 129 (90-225) | 51 (48-54) | 36 (33-40) | 708 (479-1242) | 330 (244-416) | 170 (132-210) |
| | | Non-white male | 832 (781-886) | 1218 (1139-1302) | 325 (308-342) | 120 (86-200) | 50 (48-53) | 35 (32-39) | 651 (444-1079) | 315 (233-397) | 163 (127-202) |
| | 40 | White female | 521 (500-545) | 1057 (1023-1094) | 204 (199-208) | 225 (149-452) | 52 (50-53) | 41 (39-43) | 1035 (656-2102) | 312 (228-397) | 157 (115-199) |
| | | White male | 480 (460-501) | 906 (876-938) | 187 (183-192) | 200 (138-364) | 51 (49-53) | 40 (38-42) | 906 (591-1661) | 297 (213-381) | 149 (109-191) |
| | | Non-white female | 627 (590-669) | 1208 (1134-1290) | 245 (233-258) | 208 (124-623) | 51 (48-54) | 40 (37-45) | 1024 (595-2975) | 319 (237-405) | 166 (122-210) |
| | | Non-white male | 577 (543-614) | 1035 (971-1105) | 225 (214-237) | 187 (117-478) | 51 (48-53) | 39 (36-44) | 897 (547-2253) | 305 (223-388) | 158 (117-202) |
| | 60 | White female | 362 (347-378) | 898 (866-933) | 141 (138-145) | 651 (268-NE) | 52 (50-53) | 47 (44-49) | 2631 (1061-NE) | 300 (216-384) | 150 (102-199) |

| | | | | | | | | | | | |
|---|---|---|---|---|---|---|---|---|---|---|---|
| | | White male | 333 (318-348) | 770 (742-798) | 130 (127-133) | 484 (232-NE) | 51 (50-53) | 45 (43-48) | 1917 (875-NE) | 285 (202-368) | 142 (96-188) |
| | | Non-white female | 435 (409-465) | 1027 (960-1099) | 170 (161-179) | 537 (192-NE) | 51 (48-54) | 46 (41-52) | 2293 (818-NE) | 307 (225-393) | 161 (112-211) |
| | | Non-white male | 401 (376-427) | 880 (822-941) | 156 (148-165) | 417 (176-NE) | 51 (48-54) | 45 (40-51) | 1743 (723-NE) | 293 (211-377) | 152 (106-199) |
| | 80 | White female | 251 (239-264) | 764 (728-801) | 98 (95-101) | NE (1020-NE) | 56 (52-60) | 54 (50-58) | NE (3663-NE) | 289 (203-373) | 141 (86-199) |
| | | White male | 231 (220-243) | 655 (624-685) | 90 (87-93) | NE (657-NE) | 54 (50-59) | 52 (49-57) | NE (2165-NE) | 274 (190-358) | 132 (76-186) |
| | | Non-white female | 302 (282-324) | 873 (809-942) | 118 (111-125) | NE (410-NE) | 49 (44-57) | 53 (46-63) | NE (1524-NE) | 297 (213-383) | 153 (99-210) |
| | | Non-white male | 278 (260-298) | 748 (694-805) | 109 (102-115) | NE (338-NE) | 48 (43-55) | 51 (45-61) | NE (1231-NE) | 283 (201-366) | 143 (92-200) |
| **mRNA-1273** | 20 | White female | 977 (776-1227) | | 562 (480-658) | 119 (41-NE) | | 39 (28-66) | 675 (246-NE) | | 208 (138-344) |
| | | White male | 851 (682-1071) | | 491 (421-570) | 70 (33-NE) | | 32 (24-47) | 394 (195-NE) | | 168 (115-251) |
| | | Non-white female | 912 (670-1238) | | 525 (404-684) | 231 (41-NE) | | 46 (25-266) | 1244 (248-NE) | | 239 (136-1159) |
| | | Non-white male | 797 (587-1080) | | 459 (348-598) | 98 (32-NE) | | 36 (22-110) | 527 (193-NE) | | 185 (113-477) |
| | 40 | White female | 608 (500-745) | | 351 (317-387) | NE (89-NE) | | 66 (47-111) | NE (449-NE) | | 287 (174-498) |
| | | White male | 531 (443-643) | | 306 (280-336) | 264 (60-NE) | | 48 (38-64) | 1227 (289-NE) | | 205 (130-305) |
| | | Non-white female | 569 (432-750) | | 327 (259-415) | NE (83-NE) | | 91 (38-NE) | NE (420-NE) | | 374 (162-NE) |
| | | Non-white male | 497 (374-660) | | 286 (225-363) | NE (55-NE) | | 59 (31-908) | NE (273-NE) | | 243 (125-2777) |
| | 60 | White female | 379 (294-493) | | 219 (181-265) | NE (241-NE) | | 214 (61-NE) | NE (1050-NE) | | 713 (206-NE) |
| | | White male | 331 (259-428) | | 191 (158-230) | NE (103-NE) | | 95 (46-NE) | NE (452-NE) | | 317 (141-657) |
| | | Non-white female | 354 (257-489) | | 204 (154-273) | NE (236-NE) | | 1489 (54-NE) | NE (1053-NE) | | 4634 (192-NE) |
| | | Non-white male | 310 (223-428) | | 179 (134-239) | NE (100-NE) | | 151 (40-NE) | NE (415-NE) | | 475 (139-NE) |

**Table S3. Posterior predicted peak levels (BAU/mL), half-lives (days), and time from first/second dose to the positivity threshold (days) with 95% credible intervals in participants received one vaccination with prior infection, two vaccinations without prior infection, and one vaccination without prior infection, by vaccine type**. Results were separated by age (20, 40, 60, 80-year-old), sex (female vs male) and ethnicity (white vs non-white). Estimations for two vaccinations without prior infection were based on our previous analysis[1]. NE: Not estimable, due to the antibody levels not declining in the posterior median or upper credible interval.

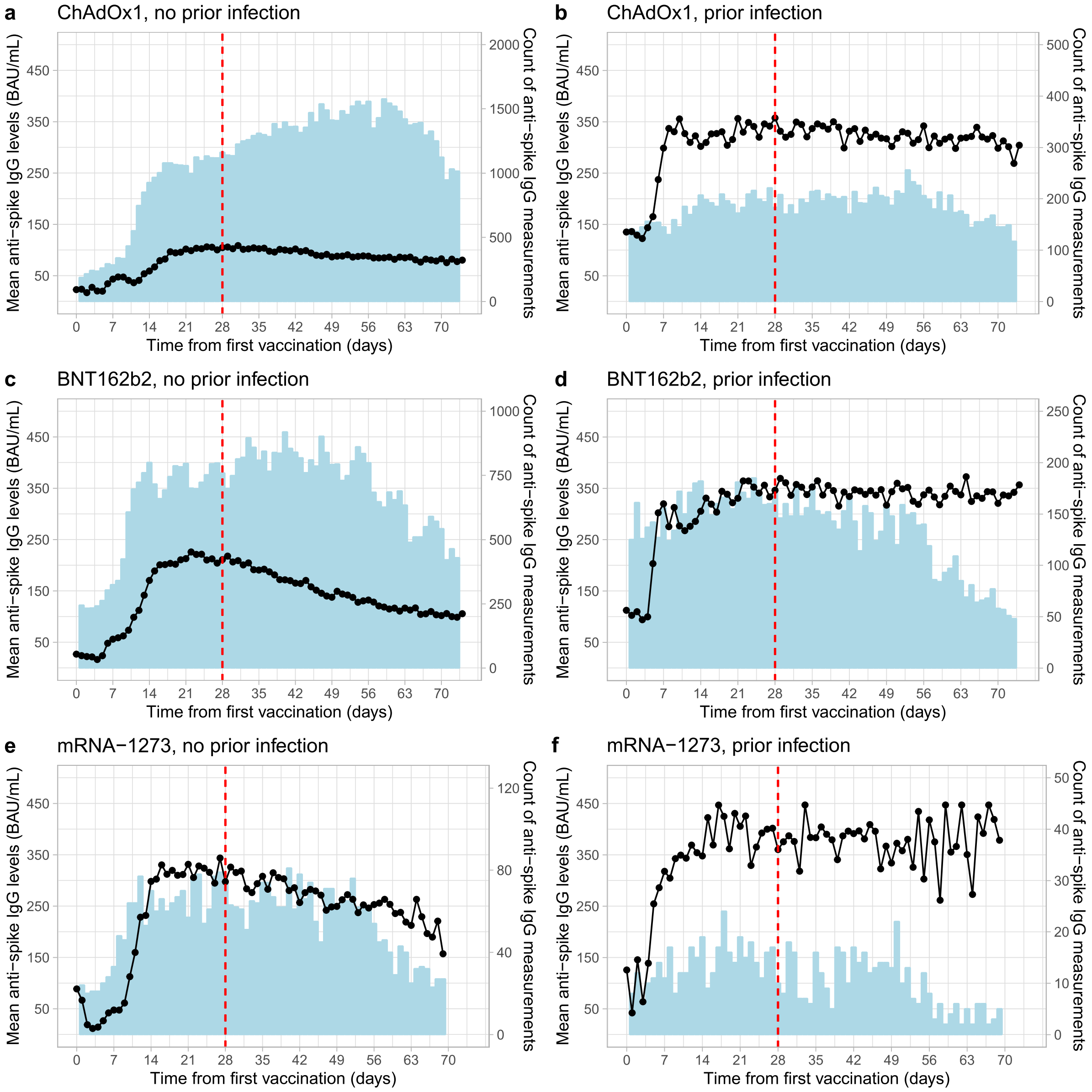


**Figure S1. Mean anti-spike IgG levels (BAU/mL) and count of anti-spike IgG measurements by day after the first vaccination.** Panels are separated by vaccines (ChAdOx1, BNT162b2, mRNA-1273) and prior infection status. Red dotted lines represent the time of the 'peak level', which is 28 days. Values truncated at 450 BAU/mL counted as =450 BAU/mL.

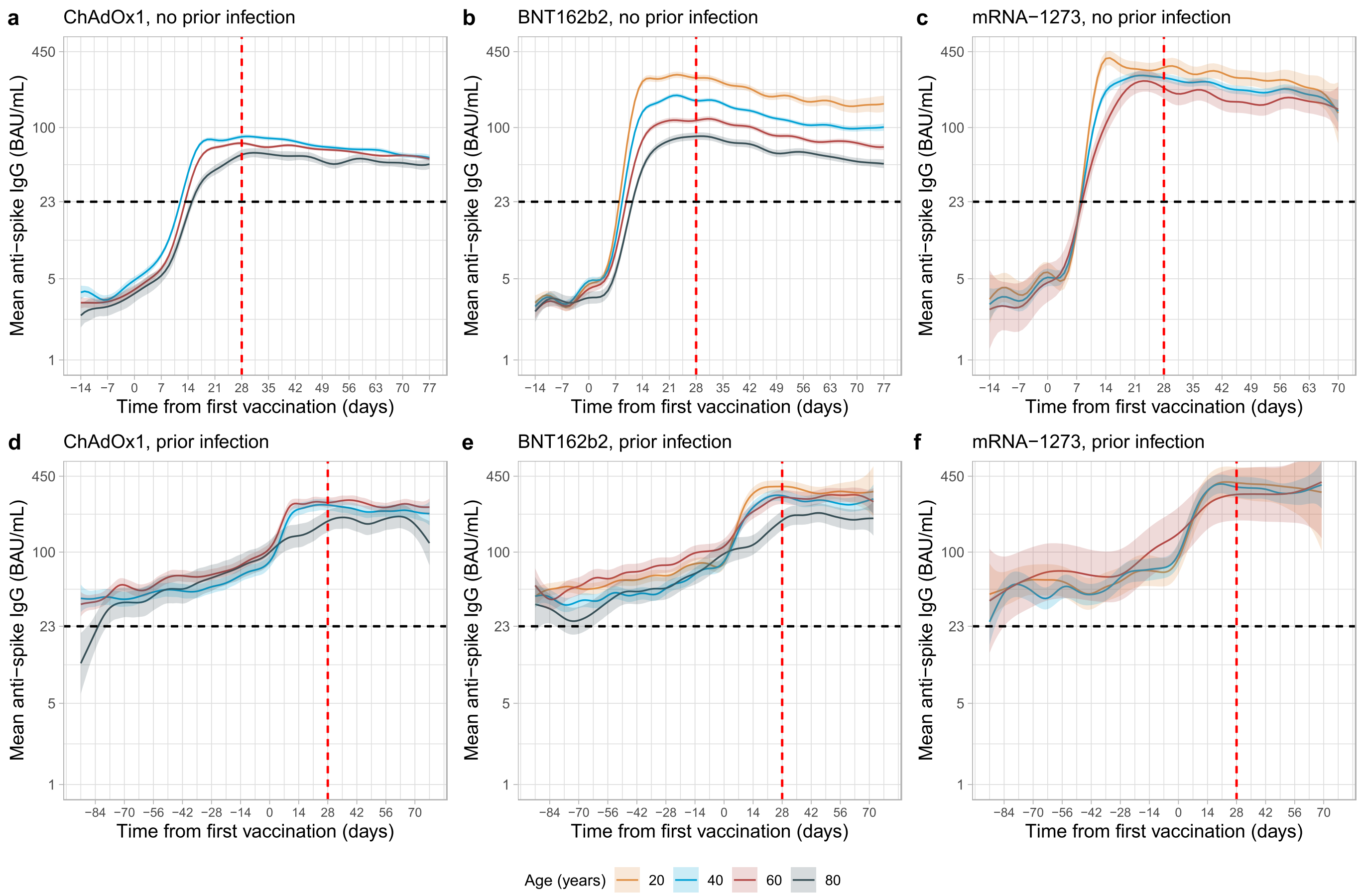


**Figure S2. Mean anti-spike IgG levels by time from first vaccination by age, vaccine type, and prior infection status. a**, No prior infection and received one ChAdOx1 vaccination, N=70,443. **b**, No prior infection and received one BNT162b2 vaccination, N=46,468. **c**, No prior infection and received one mRNA-1273 vaccination, N=3,545. **d**, With prior infection and received one ChAdOx1 vaccination, N=10,168. **e**, With prior infection and received one BNT162b2 vaccination, N=9,556. **f**, With prior infection and received one mRNA-1273 vaccination, N=779. Predicted levels are plotted on a log 10 scale. Black dotted line indicates the threshold of IgG positivity (23 BAU/mL). Red dotted lines represent the time of the 'peak level', which is 28 days. Line colour indicates response predicted for ages 20, 40, 60, and 80 years. The 95% CIs are calculated by prediction ± 1.96 × standard error of the prediction. Values truncated at 450 BAU/mL counted as =450 BAU/mL.

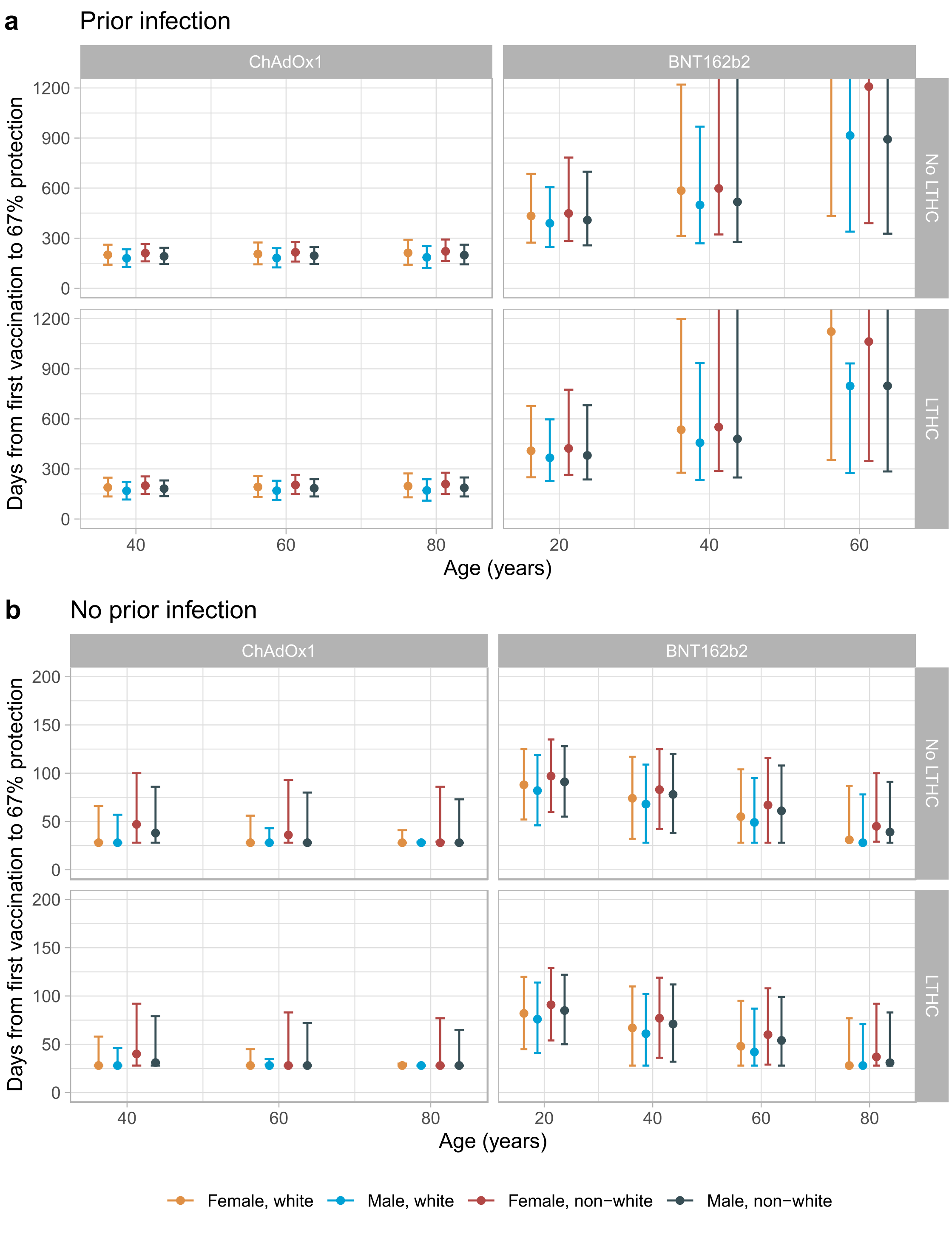


**Figure S3. Posterior predicted days (95% credible interval) from the first vaccination to the threshold level associated with 67% protection (107 BAU/mL for ChAdOx1 and 94 BAU/mL for BNT162b2) in those with evidence of prior infection (panel a) and without evidence of prior infection (panel b).** Estimates were separated by age, sex, ethnicity, long-term health condition (LTHC), and vaccine type. The correlates of protection were reported in our previous study on two vaccine doses[1]. For ChAdOx1, the 20-year-old group is not plotted because the vast majority of those receiving ChAdOx1 were ≥40 years. mRNA-1273 is not plotted because we did not have enough data to estimate its correlate of protection. In panel a, 80-year-old is not plotted in BNT162b2 because their antibody levels were not estimated to decline.
